# Decreased SARS-CoV-2 viral load following vaccination

**DOI:** 10.1101/2021.02.06.21251283

**Authors:** Matan Levine-Tiefenbrun, Idan Yelin, Rachel Katz, Esma Herzel, Ziv Golan, Licita Schreiber, Tamar Wolf, Varda Nadler, Amir Ben-Tov, Jacob Kuint, Sivan Gazit, Tal Patalon, Gabriel Chodick, Roy Kishony

**Author notes:** These authors contributed equally to this work (Matan Levine-Tiefenbrun, Idan Yelin).

## Abstract

Beyond their substantial protection of individual vaccinees, it is hoped that the COVID-19 vaccines would reduce viral load in breakthrough infections thereby further suppress onward transmission. Here, analyzing positive SARS-CoV-2 test results following inoculation with the BNT162b2 mRNA vaccine, we find that the viral load is reduced 4-fold for infections occurring 12-28 days after the first dose of vaccine. These reduced viral loads hint to lower infectiousness, further contributing to vaccine impact on virus spread.

---

The recently authorized BNT162b2 COVID-19 mRNA vaccine is about 95% efficient in preventing disease after 7 days from the second dose, and also provides some early protection starting 12 days after the first dose^1,2^. As countries race to vaccinate a significant share of the population within the coming months, the basic reproduction number of the virus is hoped to decrease. This effect can be achieved by reducing the number of susceptible people, as well as by reducing viral loads and thereby viral shedding of post-vaccination infections, turning them less infectious^3–7^. However, the effect of vaccination on viral loads in COVID-19 post-vaccination infections is yet unknown^8^.

As of January 25th, Maccabi Healthcare Services (MHS) has vaccinated over 650,000 of its members as part of a national rapid rollout of the vaccine. MHS member SARS-CoV-2 tests are often carried out in MHS central laboratory, offering the opportunity to track post-vaccination infections. Here, we retrospectively collected and analyzed the RT-qPCR test measurements of the 3 viral genes, E, N and RdRp (Allplex™ 2019-nCoV assay, SeeGene) for positive post-vaccination tests performed at MHS central laboratory between December 23rd 2020 and January 25th 2021 (n=2,897 patients, Extended Data Table 1).

Analyzing infection Ct values over time, we find that mean viral load substantially decreased 12 days post-vaccination, coinciding with the known onset of the early vaccine protection^1^. Calculating the mean Ct values for post-vaccination infections identified on each day following vaccination, we found that Ct values of positive samples collected 12-28 days after vaccination were higher than Ct values of positive samples taken during the first 11 days following vaccination (Figure 1 for RdRp, Extended Data Fig. 1 for genes N and E; Mann-Whitney U test, P<10^−8^ for the 3 genes). Differences in mean Ct calculated for these two time periods ranged from 2.1±0.2 for RdRp, through 1.9±0.2 for gene E to 1.6±0.2 for gene N.

**Figure 1.**
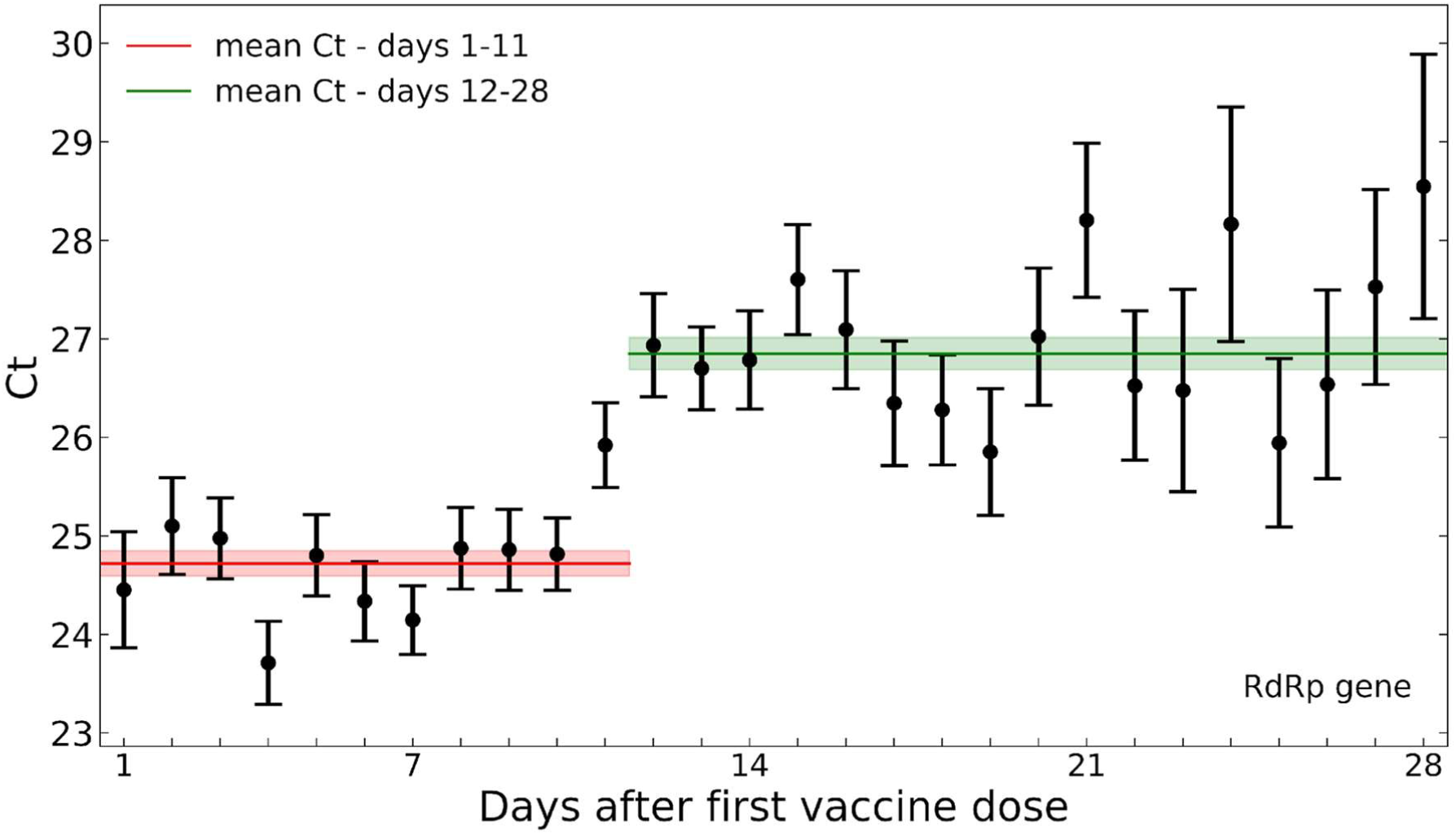
Decreased SARS-CoV-2 viral load after 12 days post-vaccination. Mean Ct values of the RdRp gene for positive tests following vaccination are plotted by the post-vaccination day in which the sample was taken. Error bars indicate the standard error of the mean. For gene E and N see Extended Data Fig. 1.

We next compared the Ct values of these post-vaccination infections with Ct values of positive tests of unvaccinated patients. As viral loads could be associated with age and sex^9^, we assembled demographically-matched control groups of positive tests in unvaccinated patients with matching age group, sex and sampling date range (Methods). Comparing post-vaccination positive tests from days 1-11 (n=1,755) with their corresponding demographically-matched control group of the same size, we found no significant difference in the distribution of Ct values for any of the 3 genes (Figure 2a for RdRp, Extended Data Fig. 2 for genes N and E). However, comparing the Ct values distribution of post-vaccination infections identified during the early protection period (days 12-28, n=1,142) with its demographically-matched unvaccinated control group of the same size, we identified a significant increase in Ct (Figure 2b for RdRp, Extended Data Fig. 2 for genes N and E; Mann Whitney U test P<10^−8^ for all 3 genes). Finally, applied on all infections (post-vaccination and unvaccinated, n=5,794), a multivariate linear regression model accounting for age, sex and vaccination quantify Ct regression coefficients ranging from 1.64 (N gene) to 2.33 (RdRp) for vaccination after 12 days or longer prior to infection sampling (Figure 2c for RdRp, Extended Data Fig. 4 for N and E genes). As a difference of 1 Ct unit is equivalent to a factor of about 1.94 in viral particles per sample^10^, these Ct differences represent a viral load ratio ranging from 2.96 to 4.68.

**Figure 2.**
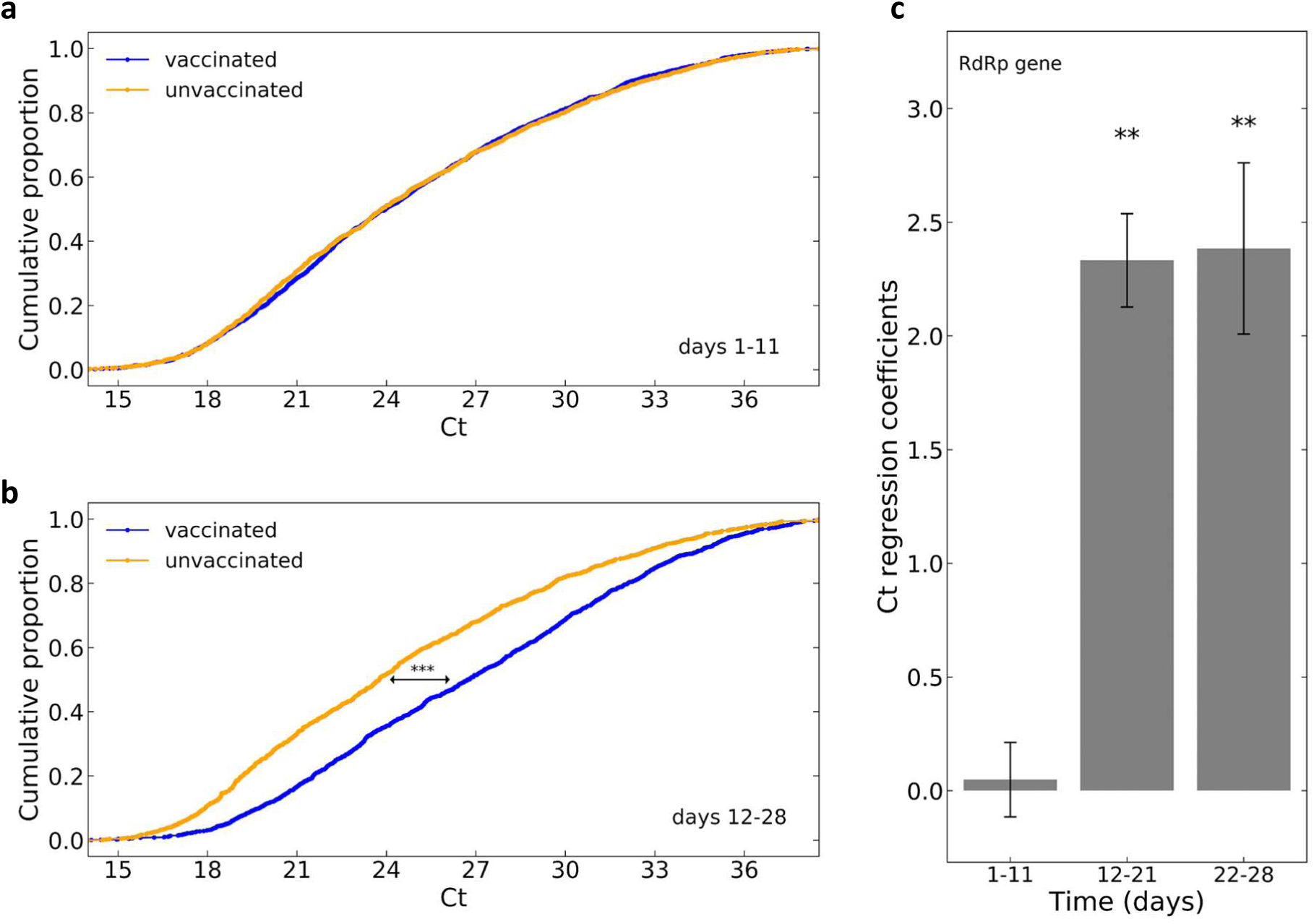
Comparison of SARS-CoV-2 viral loads among vaccinated and unvaccinated patients. **a-b**, The distribution of Ct values of the RdRp gene as determined for positive samples taken 1-11 days post-vaccination (a, n=1,755, blue) and 12-28 days post-vaccination (b, n=1,142, blue) with their respective demographically-matched control groups (orange, *** - P-value < 10^−8^, Mann-Whitney). **c**, Coefficient for the association of Ct of the RdRp gene with vaccination at different vaccination-to-sample time bins as identified by multivariate linear regression analysis accounting for age and sex (Methods). Error bars represent one standard error. For gene E and N see Extended Data Fig. 2a,b,c.

Our results show that infections occurring 12 days or longer following vaccination have significantly reduced viral loads, potentially affecting viral shedding and contagiousness as well as severity of the disease^11^. Our report is based on an observational study, not a randomized controlled trial, and has several associated limitations: (1) The group of vaccinees may differ from the demographically-matched control group in ways which could affect the observed viral load, such as behaviour, tendency to get tested, and general health status. (2) Different viral variants, which could be associated with different viral loads, may affect different parts of the population. (3) Post-vaccination positive tests may be enriched for long-term, low viral load, infections lasting from pre-immunization transmission events^9^. The average viral load may therefore further change in longer time scales post-vaccination, when infections are more strongly enriched for post-immunization transmissions. (4) The oro-nasopharyngeal test does not distinguish the viral load in the nose from the one in the oral cavity, which may be more representative of viral shedding and infectiousness. With the accumulation of additional and longer-term datasets, it will also be important to see how these results vary for other vaccines as well as among viral variants. Still, at least for the conditions tested here, the lower viral loads we observe could help tune epidemiological models of vaccine impact on the spread of the virus.

## Data Availability

To protect patient privacy, data is only available through a remote server, pending MTA.

## Acknowledgments

This work was supported by the ISRAEL SCIENCE FOUNDATION (grant No. 3633/19) within the KillCorona-Curbing Coronavirus Research Program.

## Methods

### Data collection

Anonymized SARS-CoV-2 RT-qPCR Ct values were retrieved for all positive post-vaccination samples taken between December 23rd 2020 and January 25th 2021 and tested at Maccabi Healthcare Services (MHS) central laboratory. Patients who had a positive sample prior to vaccination were excluded as well as patients age 90 and above. For patients with multiple positive post-vaccinations tests, only the first test was included. For each test, Ct values for E gene, RdRp gene, N gene and the internal control were determined using Seegene proprietary software for the Allplex™ 2019-nCoV assay following oro-nasopharyngeal swab specimens collection.

### Unvaccinated patients control group

As a control group, for each post-vaccination SARS-CoV-2 positive patient, we randomly chose an unvaccinated positive patient with similar characteristics: same sex, same age (in bins of 10 years), and same date of first positive sample (in bins of 40 days). See Extended Data Table 1.

### Linear regression

For each viral gene, linear regression of Ct values as function of Sex (0/1 Female/Male), Age (linear, in years. Adding a quadratic age term was also tested, giving very similar results), Time post vaccination (one-hot encoded binary vector for time bins 1-11, 12-21 and 22-28 days; unvaccinated encoded as all-zero). Model was implemented using Python’s statsmodels library.

### Ethics committee approval

The study protocol was approved by the ethics committee of Maccabi Healthcare Services, Tel-Aviv, Israel. IRB number: 0066-20-MHS.

## Extended data

**Extended Data Fig. 1:**
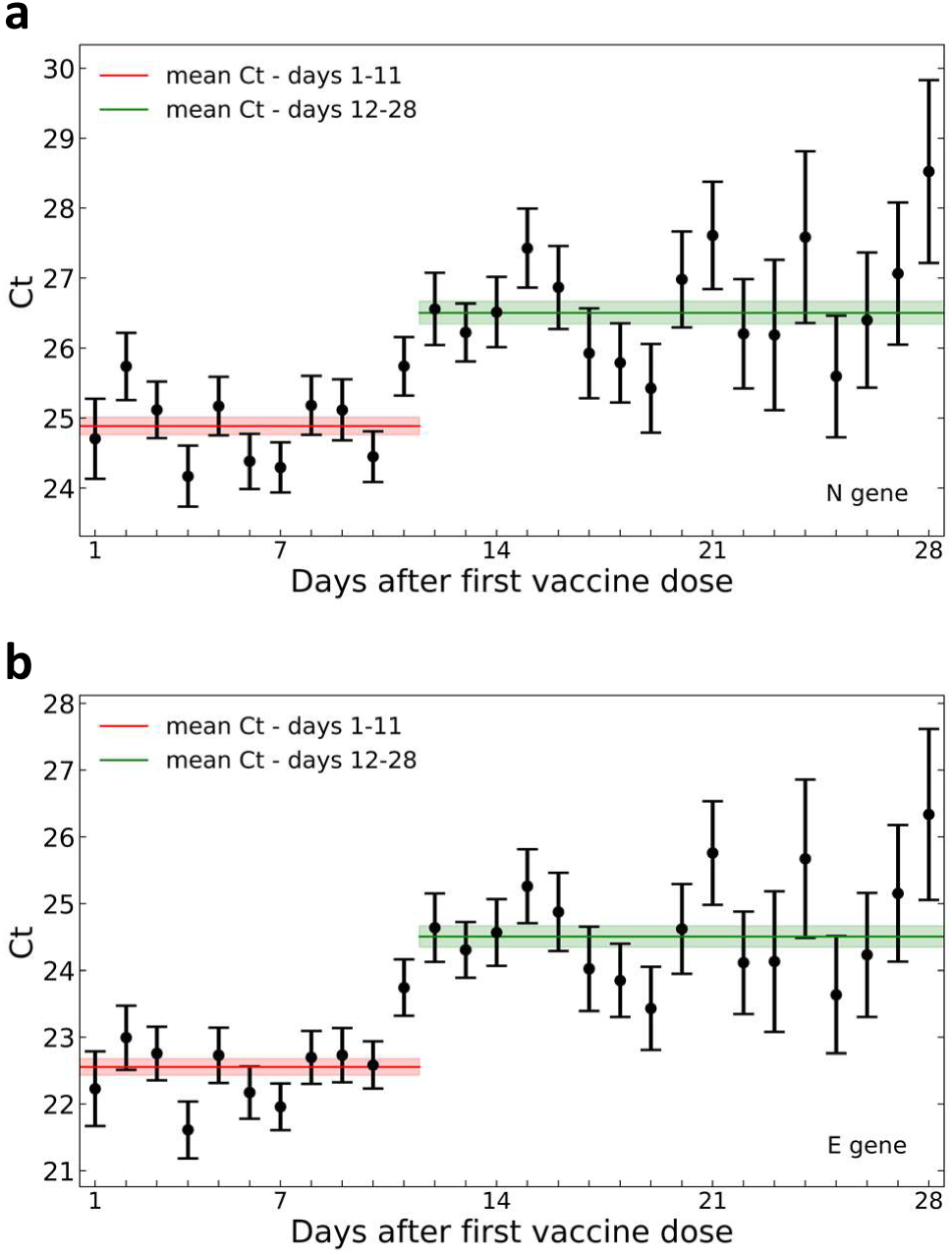
Decreased SARS-CoV-2 viral load after 12 days post-vaccination. Mean Ct values of the N and E genes for positive tests following vaccination are plotted by the day the sample was taken. Error bars indicate the standard error of the mean. **a**, N gene. **b**, E gene. For RdRp gene see Fig. 1.

**Extended Data Fig. 2:**
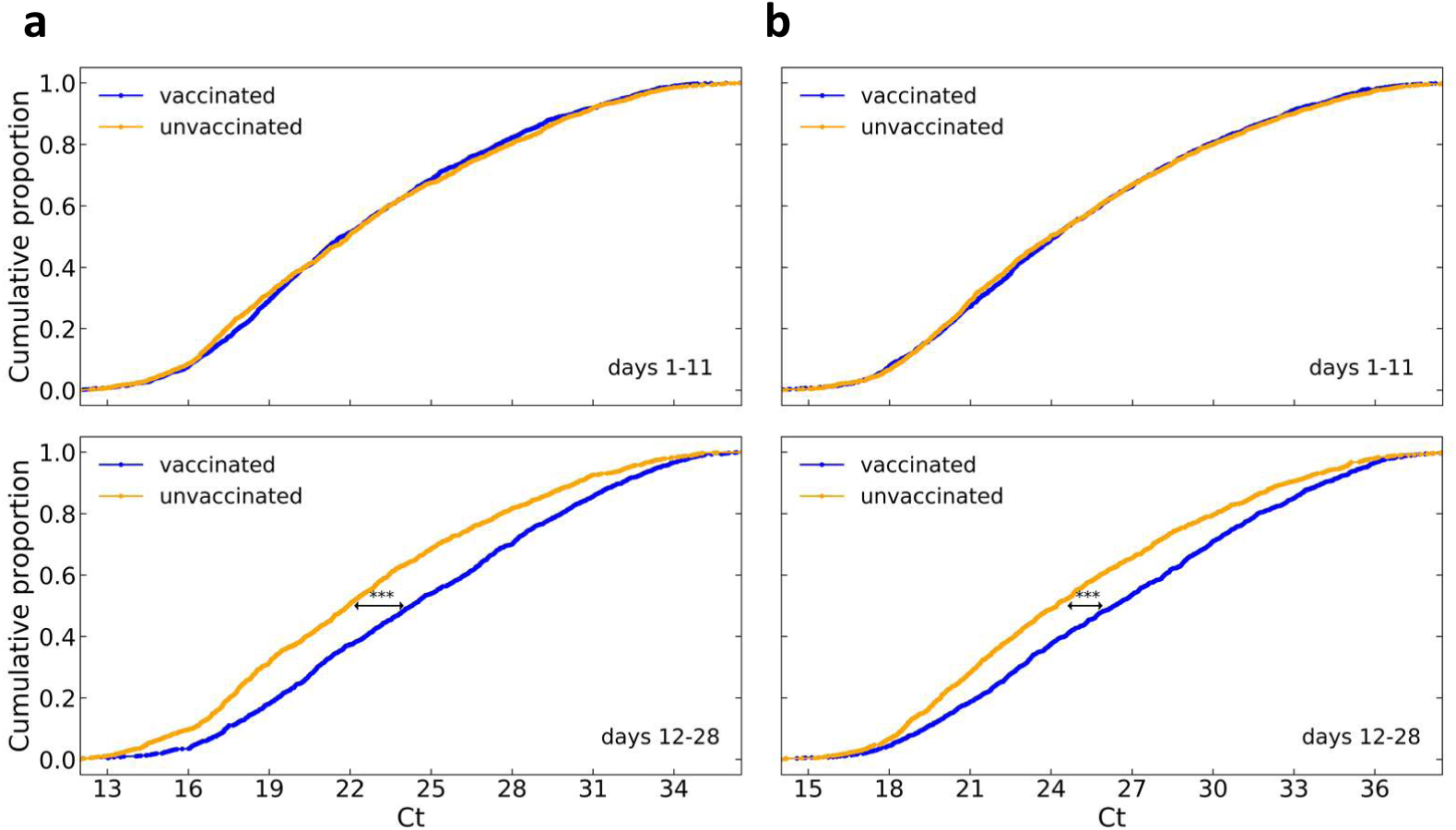
Comparison of SARS-CoV-2 viral loads among patients vaccinated prior to positive sample and unvaccinated patients. The distribution of Ct values of viral genes as determined for positive samples taken either 1-11 days post-vaccination (n=1,755, blue, top panel) or 12-28 days post vaccination (n=1,142, blue, bottom panel) with their respective demographically-matched control groups (orange). **a**, N gene. **b**, E gene. For RdRp gene see Fig. 2a.

**Extended Data Fig. 3:**
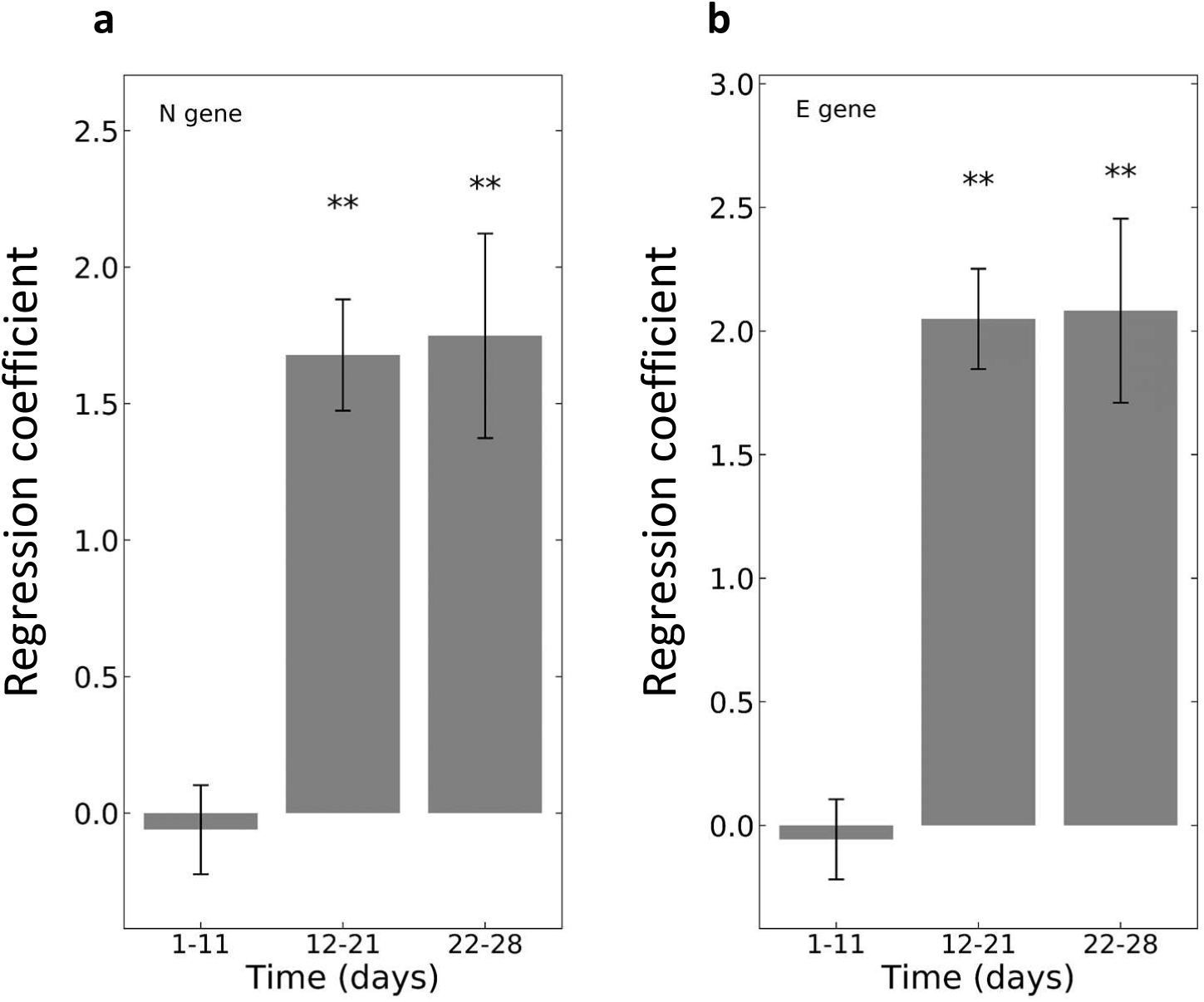
Viral load is negatively associated with vaccination starting 12 days post-vaccination. The coefficient for the association of Ct of viral genes with time of vaccination as identified by multivariate linear regression analysis accounting for age and sex (Methods). Error bars represent one standard error. **a**, N gene, **b**, E gene. For RdRp gene see Fig. 2c.

**Extended Data Table 1:** Study population.

| Age group | Total # of patients | Male | Female |
| --- | --- | --- | --- |
| 16-19 | 37 | 28 | 9 |
| 20-29 | 201 | 120 | 81 |
| 30-39 | 231 | 141 | 90 |
| 40-49 | 436 | 197 | 239 |
| 50-59 | 787 | 409 | 378 |
| 60-69 | 712 | 391 | 321 |
| 70-79 | 382 | 196 | 186 |
| 80-89 | 111 | 46 | 65 |

